# The ALDH4A1/anti-ALDH4A1 axis as a novel player of atherosclerosis in rheumatoid arthritis

**DOI:** 10.64898/2026.01.21.25342366

**Authors:** Daniel Miranda-Prieto, Mercedes Alperi-López, Ángel I. Pérez-Álvarez, Roxana Coras, Sara Alonso-Castro, Núria Amigó, Mónica Guma, Ana Suárez, Javier Rodríguez-Carrio

## Abstract

**Objectives:** Cardiovascular risk excess in rheumatoid arthritis (RA) cannot be explained by traditional risk factors alone. Recent experimental data have identified ALDH4A1 as a mitochondrial self-antigen implicated in atherosclerosis, yet its clinical significance in human autoimmunity remains unexplored. We aimed to characterize ALDH4A1 and anti-ALDH4A1 antibody levels in early RA, and evaluate their associations with atherosclerosis burden and lipoprotein traits.

**Methods:** ALDH4A1 and anti-ALDH4A1 antibodies (IgM, IgG, IgA, and IgG subclasses) were measured in early RA (n=82), clinically suspect arthralgia (n=14), healthy controls (n=70), and a validation cohort of established RA. A prospective cohort (n=13) explored therapeutic modulation under TNF blockade. Associations with atherosclerosis burden, lipid/lipoprotein profiles, oxylipin signatures, proteomics, and cell-free DNA were assessed.

**Results:** ALDH4A1 serum levels were associated with apoptotic-related proteomic pathways, cell-free DNA and lipidomic signatures in early RA. Reduced anti-ALDH4A1 antibodies were found, although divergent patters were noted across isotypes. These differences were confirmed in a validation cohort. IgG (predominantly IgG3) anti-ALDH4A1 correlated with favourable lipoprotein traits and cardiometabolic risk factors. Increased ALDH4A1 and reduced IgM/IgG anti-ALDH4A1 antibodies independently predicted atherosclerosis and improved risk stratification beyond mSCORE, most notably for IgG. ALDH4A1 tracked with TNF dynamics under TNF blockade, whereas increases in IgG antibodies occurred in responders and paralleled changes in lipoprotein features.

**Conclusions:** The ALDH4A1/anti-ALDH4A1 axis emerges as a novel player bridging lipid disturbances and atherosclerosis along the RA spectrum, hence highlighting the involvement of mitochondrial targets. These components hold promise as functional players, clinical tools and therapeutic targets.

## INTRODUCTION

Rheumatoid arthritis (RA) has been consistently associated with increased cardiovascular disease (CVD) burden, even in young patients and in the absence of traditional CV risk factors (1). Therefore, this risk excess cannot be solely explained by traditional CV risk factors, thus suggesting that non-traditional CV risk factors (2), such as autoimmunity or immune dysregulation, may play a key role.

Although both T- and B-cell responses have been implicated in atherosclerosis, the precise contribution of adaptive immunity is poorly understood. Humoral immune responses have emerged as potential drivers of atherosclerosis in both in mouse models and human studies (3). However conflicting data regarding the pathogenic or protective effect of antibodies exists (4), largely due to the poor characterization of atherosclerosis-associated antigenic targets.

Recent single-cell study provided novel insights into the germinal centre response during atherosclerosis by profiling antibody repertoires in mice using high-throughput, unbiased approaches (5). This work identified the mitochondrial enzyme ALDH4A1 as a self-antigen in the context of atherosclerosis. This protein was recognized by specific antibodies, with reactivity against atheroma and liver tissues (5). Moreover, these antibodies showed therapeutic potential in experimental studies, partly linked to restoration of lipid pathways and triglycerides in the liver (5). These findings hold promise for clinical implementation and open new avenues for understanding the interplay between immunity and metabolism in vascular disease.

Whether these findings may be applicable to other disease subsets in humans remains unknown. This question is particularly relevant in autoimmune conditions, due to the role of ALDH4A1 as self-antigen. In this regard, a deep characterization of humoral responses is needed. Furthermore, important gaps remain in the understanding of lipid metabolism during inflammation in RA (6). As a consequence, a potential role of immune responses against ALDH4A1 as a potential missing link may be hypothesized.

Therefore, the main aims of the present study were (i) to evaluate ALDH4A1 and anti-ALDH4A1 antibody levels during the early stages of RA, (ii) to assess their associations with atherosclerosis burden and evaluate their potential role as a biomarker for risk stratification, and (iii) to characterize their potential associations with lipid and lipoprotein traits in RA.

## MATERIAL AND METHODS

### Study participants

Early RA patients fulfilling the 2010 American College of Rheumatology (ACR)/European Alliance of Associations for Rheumatology (EULAR) classification criteria (7) were recruited from the early arthritic clinic of the Department of Rheumatology at Hospital Universitario Central de Asturias (HUCA). RA patients were recruited at disease onset; therefore, they were not exposed to any disease-modifying antirheumatic drugs (DMARDs) at the time of sampling. A complete medical examination including Disease Activity Score 28-joints and Health Assessment Questionnaire (HAQ) calculations was performed during the clinical appointment. Traditional CV risk factors (dyslipidaemia, hypertension, diabetes, smoking habits and obesity) were obtained from the medical records and defined in compliance with national guidelines. Individuals with clinically suspect arthralgia (CSA) (8) were also recruited from the same clinic. Moreover, healthy controls (HC) were recruited among age- and sex-matched healthy volunteers from the same population. A separate cohort of long-lasting, established RA patients recruited from the same institution were included as validation cohort (9). Additionally, a group of 13 biological-naïve RA patients (12 women, median age 43 (range: 30 – 65), DAS28 5.08(1.93), 38.5% RF+, 46.1% anti-CCP+) was prospectively followed upon TNF blockade initiation and a blood sample was drawn immediately before and 3 months after anti-TNF therapy. Clinical response after TNF blockade was evaluated according to EULAR criteria (10,11).

A fasting blood sample was collected from all individuals by venepuncture in EDTA-containing tubes. Conventional blood biochemical, including C-Reactive Protein (CRP) and Erythrocyte Sedimentation Rate (ESR) measurements, lipid analyses and complete blood counts were performed in all individuals. Blood samples were processed within less than 2 hours in the laboratory. Serum samples were stored at - 80°C until experimental procedures.

The study was approved by the local institutional review board (Comité de Ética de Investigación con Medicamentos del Principado de Asturias) in compliance with the Declaration of Helsinki (reference CEImPA 2021.126). All study subjects gave written informed consent.

### Echocardiographic study

Subclinical vascular outcomes were assessed by Doppler ultrasound at the sonograph laboratory (Department of Neurology, HUCA) in B-mode online using a Toshiba Aplio XG device (Toshiba American Medical Systems) by an experienced user, as previously described (12). In brief, the carotid intima-media thickness (cIMT) was bilaterally measured according to the “Mannheim Carotid Intima-Media Thickness Consensus (2004–2006)” (13), and atherosclerotic plaque burden (presence and extent) were defined according to established definitions as previously described (12).

Vascular stiffness was assessed by evaluating the diameter changes of the common carotid during an entire cardiac cycle, according to validated guidelines. Stiffness parameters (vascular strain (VS) and stiffness (VSf), vascular distensibility (VD) and pressure-strain elastic modulus (PSEM)) were calculated accordingly (14).

### ALDH4A1 serum levels

Levels of ALDH4A1 were quantified in serum samples using a commercial ELISA assay (P5-CDH/ALDH4A1 ELISA kit, LifeSpan BioSciences) according to the instructions provided by the manufacturer. Intra- and inter-assay coefficient of variation were <8% and <10%, respectively.

### Anti-ALDH4A1 antibody levels

Levels of anti-ALDH4A1 antibodies (IgG, IgM, IgA, and IgG subclasses: IgG1, IgG2, IgG3 and IgG4) were measured in serum samples by in-house ELISA.

ELISA MaxiSorp (Nunc) plates were coated overnight with 2,5 µg/ml ALDH4A1 (MedChemExpress) in carbonate buffer (pH=9.6) at room temperature. The plates were blocked with 1% bovine serum albumin (BSA) in phosphate buffer saline (PBS) (pH 7.4) for 1 hour at 37° C and then washed with PBS. Next, standard curves (1:8 to 1:256 diluted pooled sera), diluted samples (IgG: 1:25, IgM: 1:10, IgA: 1:10, IgG subclasses: 1:10) were prepared in 0.1% BSA PBS and incubated for 2 h at room temperature under shaking conditions. Unbound antibodies were removed by repeating the washing step. Alkaline phosphatase-conjugated anti-human IgG (1:1000), IgM (1:500) (both from Immunostep), IgA (1:800) (Abcam antibodies) in 1% BSA tris buffer saline (TBS) was added and incubated for 1h. Finally, the plate was washed twice with TBS and 1 mg/ml p-nitrophenylphosphate (Sigma) diluted in dietanolamine buffer (pH 9.8) was added to the wells. Absorbance at 405 nm was recorded and anti-ALDH4A1 Arbitrary Units (AU) were calculated for each sample according to the standard curves. Intra- and inter-assay coefficient of variation ranges were (4.2-7.8%) and (8.1-11.5%) for all the assays, respectively.

### Proinflammatory cytokines

IFNa serum levels were quantified using a Cytometric Bead Array Flex Set (BD). The serum levels of IL-6, TNF, IFNg, IL-1b, IL-23, IL-12, IL-33, IL-10, IL-17, IL-8, and IL-18 were measured by a predefined multiplex assay (Human Inflammation Panel 1, LEGENDplex, BioLegend). APRIL and BAFF serum levels were quantified using ELISA kits (Invitrogen, ThermoFisher) in accordance with the manufacturer’s instructions.

### Lipoprotein characterization

Lipoprotein profiles were characterized by means of the H-NMR-based Liposcale test (15). This validated approach allowed the assessment of lipid content (cholesterol and triglycerides) of the major lipoprotein classes (VLDL, IDL, LDL and HDL) as well as the particle number and size (diameter) of VLDL LDL and HDL and their subclasses (small, medium and large).

In brief, serum samples were diluted with 50 mM pH 7.4 phosphate buffer (with deuterated water) and undergone H-NMR analysis. Then, H-NMR spectra were recorded on a Bruker Advance III 600 spectrometer at 310 K operating at a proton frequency of 600.20 MHz. Particle concentration and the diffusion coefficients were obtained from the measured amplitudes and attenuation of their spectroscopically distinct lipid methyl group NMR signals, using the 2D diffusion-ordered 1H NMR spectroscopy (DSTE) pulse as previously described (15).

### Eicosanoid-derived oxylipins analysis

Eicosanoid-derived oxylipins levels were evaluated in serum samples by LC/MS-MS as previously described (16). In brief, samples were spiked and purified by solid phase extraction on Strata-X columns. Then, oxylipins were separated by reverse-phase chromatography under an Acquity UPLC system (Waters). The LC was interfaced with an IonDrive Turbo V ion source, and mass spectral analysis was performed on a triple quadrupole AB SCIEX 6500 QTrap mass spectrometer (AB SCIEX, Framingham, MA). Oxylipins were measured using electrospray ionization in negative ion mode and multiple reaction monitoring (MRM) using the most abundant and specific precursor ion/product ion transitions to build an acquisition method capable of detecting 158 analytes and 26 internal standards, as previously described (17,18). Oxylipins were identified by matching their MRM signal and chromatographic retention time with those of pure identical standards. Finally, oxylipins and free fatty acids were quantified by the stable isotope dilution method, and levels were shown as pmol/ml.

### Proteomic platform

Serum proteomics were evaluated by means of a high-throughput analysis. A predefined panel of 92 protein hits related to CVD (Cardiovascular Panel II) was assessed using the Proximity Extension Assay provided by Olink (Olink Bioscience, Sweden) (19).

### Cell-free DNA

Platelet-poor plasma (PPP) was prepared from blood samples obtained in tubes containing citrate-containing tubes from each participant and stored at -80°C within less than 2 hours from collection.

Total circulating DNA (cirDNA) was isolated from PPP via a magnetic bead-based extraction method (MagMAX Cell-free DNA Isolation Kit, Applied Biosystems, Massachusetts, United States), as previously described (20). Then, cirDNA was quantified in a Qubit fluorometer using the Qubit dsDNA HS Assay Kit (Thermo Fisher).

Next, mitochondrial and nuclear cell-free DNA (mtDNA and nDNA, respectively) were analysed and quantified from previously isolated cirDNA by means of a quantitative real-time PCR (qPCR) analysis using SYBR Green and specific primer sets, as previously described (20).

### Statistical analyses

Variables were summarized as median (interquartile range) or mean±standard deviation, or n(%), as appropriate.

Differences were evaluated by Mann-Withney U, Kruskal-Wallis tests with Dunn-Bonferroni correction for multiple comparisons, or Wilcoxon paired tests. Correlations were assessed by Spearman ranks’ tests. The associations between ALDH4A1 or antibody levels and atherosclerosis occurrence was analysed by multiple logistic regression adjusted by confounders. Odds ratio (OR) and 95% confidence intervals (CI) were calculated. In order to add ALDH4A1 or antibody levels to the mSCORE algorithm, categories were defined from their distribution in the HC population using tertiles as cut-offs, and scores (0, 1 or 2) were given accordingly, as described elsewhere (12).

The performance of the risk reclassification was evaluated by classification measures (sensitivity, specificity, accuracy), discriminative effect (AUC ROC), Matthews’ correlation coefficient, goodness of fit (Hosmer–Lemeshow test) and the Youden index to determine the optimal cut-offs maximizing accuracy. Differences in the ROC values were compared by the De Jong statistic. Net Reclassification Improvement (NRI) was used to compared among modified models.

Proteomic data were evaluated by pathway enrichment analyses using the STRING database and KEGG platforms (21,22). For eicosanoid-derived oxylipins, pathway analyses were performed using RaMP-DB database under MetaboAnalyst v.6.0 (23), and Raw p, (adjusted) Holm p and enrichment ratios were obtained for each pathway.

A p-value <0.050 was considered as statistically significant. Statistical analyses were carried out under SPSS v. 27, R v.4.1.3 and GraphPad Prism 8.0.

## RESULTS

### 1. ALDH4A1 and anti-ALDH4A1 antibody levels in RA

A total of 82 RA patients, 14 individuals with CSA and 70 HC were recruited for this study (Table 1). No differences in ALDH4A1 serum levels were found among early RA, CSA and HC (Figure 1A). Equivalent results were obtained in established RA (Supplementary Table 1), as a validation cohort (Figure 1B). Proteomic analyses revealed that ALDH4A1 was associated with 32 protein hits in early RA, with a significant protein-protein interaction enrichment (p=5.8·10^-8^) network (Figure 1C). Pathway annotation informed functional pathways related to apoptosis, TNF-related signalling, and osteoclast differentiation (Figure 1D). Furthermore, ALDH4A1 was associated with cfDNA levels (r=0.378, p<0.001) and with nDNA (r=0.305, p=0.005), but not mtDNA (r=0.103, p=0.358), in early RA.

**Figure 1:**
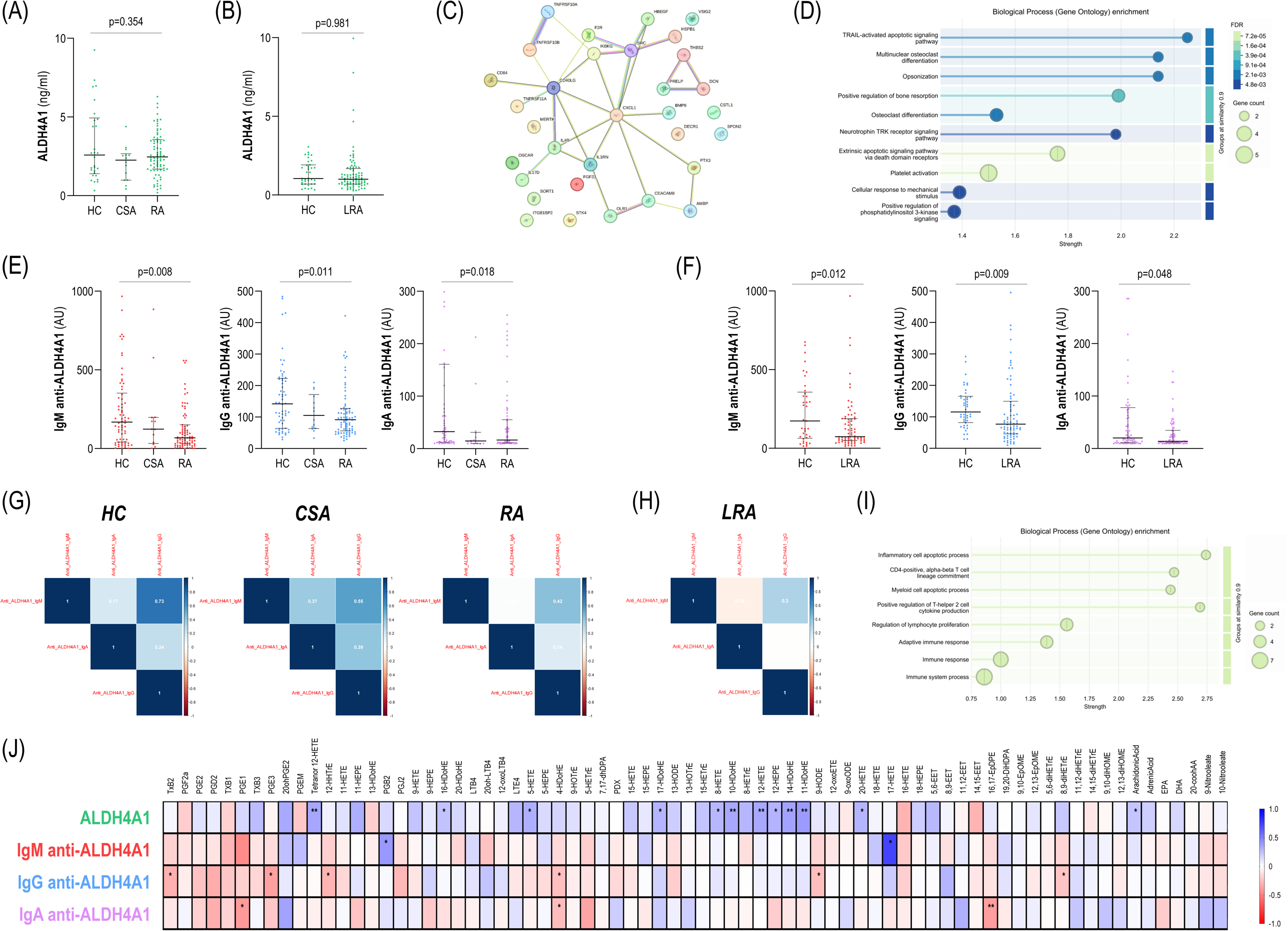
Assessment of ALDH4A1 and anti-ALDH4A1 serum levels in RA. (A) ALDH4A1 serum levels in HC, CSA and RA, as well as in HC and LRA groups (B) (validation cohort). (C) Protein-protein interactions among protein hits found to be associated with ALDH4A1 serum levels in early RA. (C) Pathway annotation (top 10) retrieved by the STRING platform using GO enrichment. Pathways were ranked by strength, and both FDR and gene count are indicated. (E) Anti-ALDH4A1 antibody (IgM, IgG and IgA) serum levels in HC, CSA and RA, as well as in HC and LRA groups (F) (validation cohort). Each dot represents one individual. Bars represent 25th percentile (lower), median and 75th percentile (upper). Differences were assessed by Kruskal-Wallis (with Dunn-Bonferroni correction for multiple comparisons) or Mann-Withney U tests, as appropriate. P-values are indicated. (G) The correlations among different isotypes were studied across study groups in correlograms (G-H). Correlation coefficients for each pair of variables are shown (white). Colour gradient varied from blue (positive correlations) to red (negative correlations) according to scale (right). (I) Pathway annotation (top 10) retrieved by the STRING platform using GO enrichment from protein hits found to be associated with IgG anti-ALDH4A serum levels. Pathways were ranked by strength, and both FDR and gene count are indicated. (I) Matrix depicting the correlations between eicosanoid-derived oxylipin species (columns) and ALDH4A1 or anti-ALDHA1 antibody levels (rows). Each tile represents a correlation coefficient. Colour gradient varied from blue (positive correlations) to red (negative correlations) according to scale (right). The statistically significant p-values from the correlation analyses are indicated as follows: * p<0.050, and ** p<0.010.

**Table 1:** Demographic and clinical features of study participants. Demographic and clinical features of study participants were summarized. Variables were expressed as median (interquartile range) or n(%), unless otherwise stated, according to the distribution of the variables.

|  | **HC**<br>**n=70** | **CSA**<br>**n=14** | **RA**<br>**n=82** |
| --- | --- | --- | --- |
| Age (years), mean (range) | 56.40<br>(35.10-79.30) | 49.28<br>(30.00-62.33) | 58.51<br>(30.33-83.67) |
| Gender (female/male) | 28/18 | 14/0 | 66/16 |
| ***Clinical features*** |  |  |  |
| Duration of symptoms (weeks) |  | 24.00 (40.00) | 20.00 (22.00) |
| Morning stiffness (minutes) |  | 30.00 (50.00) | 60.00 (80.0) |
| Tender joint count |  | 3.00 (3.00) | 8.00 (7.00) |
| Swollen joint count |  | 0.00 (1.00) | 6.00 (5.00) |
| Patient global assessment<br>(VAS 0-100) |  | 30.00 (50.00) | 70.00 (25.00) |
| DAS28 |  |  | 5.40 (1.78) |
| HAQ |  | 0.55 (0.60) | 1.11 (1.00) |
| Pain (VAS 0-10) |  | 5.00 (5.00) | 7.00 (2.00) |
| RF+, n(%) |  | 8 (66.6) | 57 (69.5) |
| ACPA+, n(%) |  | 7 (58.3) | 56 (68.2) |
| ***Laboratory parameters*** |  |  |  |
| ESR (mm/h) |  | 7.50 (8.84) | 24.00 (27.00) |
| CRP (mg/dl) |  | 0.15 (0.30) | 0.80 (2.20) |
| C3 (g/l) |  | 1.23 (0.34) | 1.33 (0.42) |
| C4 (g/l) |  | 0.22 (0.08) | 0.29 (0.12) |
| Glucose (mg/dl) |  | 96.00 (21.00) | 92.00 (10.00) |
| Urate (mg/dl) |  | 3.85 (0.70) | 4.60 (1.70) |
| Alanine aminotransferase<br>(AAT) (U/l) |  | 13.50 (8.25) | 17.00 (11.00) |
| ***Traditional CV risk factors,***<br>***n(%)*** |  |  |  |
| Hypertension |  | 1 (12.5) | 28 (34.1) |
| Diabetes |  | 0 (0.0) | 9 (10.9) |
| Dyslipidemia |  | 3 (37.5) | 24 (29.2) |
| Smoking |  | 10 (71.4) | 31 (27.8) |
| Obesity (BMI>30 kg/m<sup>2</sup>) |  | 3 (37.5) | 33 (40.2) |

Decreased levels of anti-ALDH4A1 antibodies were observed across isotypes (IgM, IgG, and IgA) in early RA patients compared to HC (Figure 1E). Interestingly, IgM and IgG levels in the CSA group lied between those of RA and HC. These findings were confirmed in the validation cohort (Figure 1F). Further analyses revealed that IgG responses were mostly predominated by IgG3 isotype, and IgG3 anti-ALDH4A1 antibodies recapitulated differences among study groups (Supplementary Figure 1). Correlation analyses revealed strong associations among isotypes (namely IgG and IgM) in HC, whereas a progressive decline was noted towards CSA and RA groups (Figure 1G). Results from the validation cohort supported this observation (Figure 1H). Proteomic correlates were minimal for IgM (4) and IgA (2), whereas 9 protein hits were retrieved for the IgG isotype. Protein network (PPI enrichment p<0.0001) allowed the identification of functional pathways related to overall immune activation and Th2 regulation (Figure 1I). No associations between anti-ALDH4A1 levels and cfDNA (either nDNA or mtDNA) were found (all p>0.050).

ALDH4A1 and anti-ALDH4A1 serum levels were unrelated to clinical features and traditional risk factors in early RA patients (Supplementary Table 2), except for certain traditional CV risk factors (dyslipidaemia and obesity) and biochemical parameters (urate, AAT, C3 and C4 levels) for IgG. Moreover, these correlations were stronger with IgG3 anti-ALDH4A1 levels, whereas no associations were retrieved with other subclasses. No associations with proinflammatory cytokines were detected overall (Supplementary Table 3) although, IgG-to-IgM anti-ALDH4A1 ratio was positively correlated to APRIL levels (r=0.378, p=0.030) in RA. Interestingly, IgG anti-ALDH4A1 antibodies were found to be associated with lipoprotein disturbances in early RA, whereas no associations were observed for the rest of isotypes or ALDH4A1 serum levels (Table 2). Further analyses revealed associations between ALDH4A1 levels with a number of eicosanoid-derived oxylipin species (n=12) in early RA (Figure 1J). Pathway analyses confirmed an enrichment of arachidonic acid metabolism, followed by alpha-linolenic, eicosanoid and docosahexaenoic metabolic pathways (Supplementary Table 4). A distinct profile of associations was noticed for anti-ALDH4A1 antibodies (IgM: n=2, IgG: n=6, and IgA: n=3), although associations were overall modest (Figure 1 J).

**Table 2:** Associations between ALDH4A1 and anti-ALDH4A1 antibodies and lipoprotein traits. Associations between levels of ALDH4A1 and anti-ALDH4A1 antibodies and lipoprotein features (particle content, particle size and subclasses) were analysed by Spearman’s rank tests in RA and CSA groups. Correlation coefficients (r) and p-values are shown. Those reaching statistical significance are highlighted in bold.

|  | RA |  |  |  | CSA |  |  |  |
| --- | --- | --- | --- | --- | --- | --- | --- | --- |
|  | ALDH4A1 | IgM anti-ALDH4A1 | IgG anti-ALDH4A1 | IgA anti-ALDH4A1 | ALDH4A1 | IgM anti-ALDH4A1 | IgG anti-ALDH4A1 | IgA anti-ALDH4A1 |
| *Particle content* |  |  |  |  |  |  |  |  |
| VLDL-C | r=0.102<br>p=0.361 | r=0.002<br>p=0.983 | r=-0.142<br>p=0.202 | r=-0.061<br>p=0.587 | r=0.271<br>p=0.349 | **r=-0.547**<br>**p=0.043** | r=-0.240<br>p=0.409 | r=-0.097<br>p=0.742 |
| IDL-C | r=0.159<br>p=0.155 | r=-0.111<br>p=0.322 | r=-0.188<br>p=0.091 | r=-0.137<br>p=0.220 | r=0.442<br>p=0.113 | r=-0.490<br>p=0.075 | r=-0.174<br>p=0.553 | r=0.033<br>p=0.911 |
| LDL-C | r=-0.114<br>p=0.309 | r=-0.126<br>p=0.260 | **r=-0.346**<br>**p=0.001** | r=0.096<br>p=0.390 | r=0.240<br>p=0.409 | r=-0.244<br>p=0.401 | r=0.130<br>p=0.659 | r=-0.066<br>p=0.823 |
| HDL-C | r=0.001<br>p=0.994 | r=-0.122<br>p=0.273 | r=-0.151<br>p=0.176 | r=0.159<br>p=0.154 | r=-0.310<br>p=0.280 | r=0.437<br>p=0.118 | r=0.218<br>p=0.455 | r=0.031<br>p=0.917 |
| VLDL-TG | r=0.099<br>p=0.376 | r=-0.090<br>p=0.422 | **r=-0.229**<br>**p=0.039** | r=-0.116<br>p=0.299 | r=0.403<br>p=0.153 | **r=-0.565**<br>**p=0.035** | r=-0.319<br>p=0.267 | r=-0.273<br>p=0.345 |
| IDL-TG | **r=0.230**<br>**p=0.038** | r=-0.094<br>p=0.403 | r=-0.171<br>p=0.124 | r=-0.115<br>p=0.304 | r=0.460<br>p=0.098 | r=-0.481<br>p=0.081 | r=-0.200<br>p=0.493 | r=-0.026<br>p=0.929 |
| LDL-TG | r=0.058<br>p=0.603 | r=-0.098<br>p=0.383 | r=-0.169<br>p=0.128 | r=-0.007<br>p=0.953 | r=0.244<br>p=0.400 | r=-0.332<br>p=0.246 | r=-0.077<br>p=0.794 | r=0.086<br>p=0.771 |
| HDL-TG | r=0.208<br>p=0.060 | r=0.035<br>p=0.757 | r=0.126<br>p=0.257 | r=-0.075<br>p=0.500 | r=0.262<br>p=0.366 | r=-0.187<br>p=0.523 | r=0.037<br>p=0.899 | r=-0.004<br>p=0.988 |
| **Particle number** |  |  |  |  |  |  |  |  |
| VLDL-P | r=0.114<br>p=0.306 | r=-0.071<br>p=0.524 | **r=-0.225**<br>**p=0.042** | r=-0.104<br>p=0.354 | r=0.304<br>p=0.291 | r=-0.512<br>p=0.061 | r=-0.275<br>p=0.342 | r=-0.189<br>p=0.517 |
| Large | r=0.105<br>p=0.349 | r=-0.043<br>p=0.704 | r=-0.145<br>p=0.194 | r=-0.072<br>p=0.520 | r=0.332<br>p=0.246 | **r=-0.578**<br>**p=0.030** | r=-0.358<br>p=0.208 | r=-0.233<br>p=0.422 |
| Medium | r=0.033<br>p=0.767 | r=-0.036<br>p=0.746 | r=-0.141<br>p=0.208 | r=-0.119<br>p=0.287 | r=0.244<br>p=0.400 | r=-0.415<br>p=0.140 | r=0.007<br>p=0.982 | r=-0.090<br>p=0.759 |
| Small | r=0.121<br>p=0.280 | r=-0.067<br>p=0.552 | **r=-0.228**<br>**p=0.039** | r=-0.104<br>p=0.350 | r=0.299<br>p=0.299 | r=-0.499<br>p=0.069 | r=-0.301<br>p=0.296 | r=-0.282<br>p=0.329 |
| LDL-P | r=-0.068<br>p=0.546 | r=-0.149<br>p=0.182 | **r=-0.391**<br>**p<0.001** | r=0.072<br>p=0.523 | r=0.306<br>p=0.288 | r=-0.429<br>p=0.126 | r=-0.064<br>p=0.829 | r=-0.242<br>p=0.404 |
| Large | r=-0.136<br>p=0.222 | r=0.018<br>p=0.869 | r=-0.086<br>p=0.441 | r=0.021<br>p=0.852 | r=0.139<br>p=0.637 | r=0.081<br>p=0.782 | r=0.521<br>p=0.056 | r=0.216<br>p=0.459 |
| Medium | r=-0.079<br>p=0.479 | r=-0.101<br>p=0.366 | r=-0.190<br>p=0.087 | r=0.064<br>p=0.565 | r=0.271<br>p=0.349 | r=-0.204<br>p=0.483 | r=0.262<br>p=0.366 | r=0.048<br>p=0.869 |
| Small | r=-0.039<br>p=0.730 | r=-0.168<br>p=0.132 | **r=-0.509**<br>**p<0.001** | r=0.047<br>p=0.674 | r=0.411<br>p=0.144 | **r=-0.578**<br>**p=0.030** | r=-0.209<br>p=0.474 | r=-0.288<br>p=0.318 |
| HDL-P | r=0.018<br>p=0.871 | r=-0.147<br>p=0.189 | r=-0.201<br>p=0.071 | r=0.147<br>p=0.189 | r=-0.110<br>p=0.708 | r=0.213<br>p=0.464 | r=0.138<br>p=0.637 | r=0.136<br>p=0.642 |
| Large | r=-0.018<br>p=0.875 | r=-0.026<br>p=0.818 | r=-0.130<br>p=0.244 | r=0.018<br>p=0.869 | r=-0.174<br>p=0.552 | r=0.029<br>p=0.923 | r=0.095<br>p=0.748 | r=0.059<br>p=0.840 |
| Medium | r=0.070<br>p=0.531 | r=0.041<br>p=0.712 | r=0.110<br>p=0.323 | r=0.053<br>p=0.637 | r=-0.513<br>p=0.061 | **r=0.565**<br>**p=0.035** | r=0.253<br>p=0.383 | r=0.154<br>p=0.599 |
| Small | r=0.025<br>p=0.821 | r=-0.190<br>p=0.088 | **r=-0.290**<br>**p=0.008** | r=0.145<br>p=0.195 | r=0.156<br>p=0.594 | r=-0.055<br>p=0.852 | r=-0.059<br>p=0.840 | r=0.048<br>p=0.869 |

| *Particle diameter* |  |  |  |  |  |  |  |  |
| --- | --- | --- | --- | --- | --- | --- | --- | --- |
| VLDL | r=-0.122<br>p=0.276 | r=0.097<br>p=0.388 | **r=0.241**<br>**p=0.029** | r=0.086<br>p=0.440 | r=-0.374<br>p=0.188 | r=0.499<br>p=0.069 | r=0.349<br>p=0.221 | r=0.286<br>p=0.322 |
| LDL | r=-0.088<br>p=0.434 | r=0.135<br>p=0.227 | **r=0.258**<br>**p=0.019** | r=-0.023<br>p=0.837 | r=-0.356<br>p=0.211 | r=0.512<br>p=0.061 | **r=0.565**<br>**p=0.035** | r=0.420<br>p=0.135 |
| HDL | r=-0.036<br>p=0.746 | r=0.106<br>p=0.343 | **r=0.359**<br>**p<0.001** | r=-0.097<br>p=0.386 | r=-0.337<br>p=0.089 | r=0.262<br>p=0.366 | r=0.059<br>p=0.840 | r=0.020<br>p=0.946 |

**Table 3:** ALDH4A1 and anti-ALDH4A1 antibodies as predictors of atherosclerosis. The role of ALDH4A1 or anti-ALDH4A1 antibody levels as predictors of atherosclerosis occurrence was analyzed by univariate and multivariate logistic regression models in RA patients. Models were run independently for each predictor. Model 1 represents a univariate model. Model 2 represents a multivariate model adjusted for traditional CV risk factors (age, sex, hypertension, dyslipidaemia, diabetes, smoking, and obesity). Model 3 represents a multivariate model adjusted for traditional CV risk factors and clinical features (model 2 + disease activity, RF, and ACPA positivity). Odds ratios (OR), 95% confidence intervals and p-values were calculated for each predictor and model. Those reaching statistical significance were highlighted in bold.

|  | Model 1 |  | Model 2 |  | Model 3 |  |
| --- | --- | --- | --- | --- | --- | --- |
|  | *OR [95% CI]* | *p-value* | *OR [95% CI]* | *p-value* | *OR [95% CI]* | *p-value* |
| ALDH4A1 | 1.805 [1.195 – 2.727] | **0.012** | 1.678 [1.015 – 2.772] | **0.043** | 1.855 [1.004 – 3.425] | **0.045** |
| IgM anti-ALDH4A1 | 0.898 [0.827 – 0.976] | **0.011** | 0.840 [0.736 – 0.959] | **0.010** | 0.846 [0.734 – 0.970] | **0.022** |
| IgG anti-ALDH4A1 | 0.838 [0.715 – 0.982] | **0.017** | 0.722 [0.565 – 0.923] | **0.006** | 0.718 [0.540 – 0.945] | **0.013** |
| IgA anti-ALDH4A1 | 1.022 [0.940 – 1.111] | 0.611 | 0.998 [0.885 – 1.125] | 0.971 | 0.977 [0.856 – 1.115] | 0.726 |

Altogether, these findings point to alterations in the ALDH4A1/anti-ALDH4A1 axis along the whole RA spectrum, especially in the early stage. ALDH4A1 may be related to specific proteomic and lipidomic signatures, while divergent profiles were found within anti-ALDH4A1 responses.

### 2. ALDH4A1/anti-ALDH4A1 axis as a biomarker of atherosclerosis in early RA

Next, the associations between ALDH4A1 and anti-ALDH4A1 antibodies and atherosclerosis (Supplementary Table 5), alone or in combination with traditional CV risk factors, were analysed.

Atherosclerosis occurrence was linked to increased ALDH4A1 and decreased IgM/IgG anti-ALDH4A1 levels in early RA, whereas no differences in IgA anti-ALDH4A1 were found (Figure 2A). Similar results were observed for plaque extent (Supplementary Table 6). ALDH4A1 was also associated with atherosclerosis in CSA (Supplementary Table 6). Interestingly, analysis of the IgG3 subclass mirrored the differences observed for the whole IgG isotype (Supplementary Figure 2), hence strengthening previous findings. ALDH4A1, IgM and IgG anti-ALDH4A1 levels were independently associated with atherosclerosis occurrence in univariate and multivariate models after adjusted for traditional risk factors (Table 2), whereas no effect was registered for their IgA counterparts. Adjustment for clinical features did not change these findings (Table 2). A similar picture was obtained for plaque extent (number of plaques) prediction using Poisson log-lineal models (ALDH4A1: 1.113 [1.013-1.222], p=0.026; and IgG anti-ALDH4A1: 0.900 [0.821-0.987], p=0.019). The analysis of history of CV events in the validation cohort confirmed these findings (ALDH4A1: p=0.007, IgM: p=0.012; IgG anti-ALDH4A1: p=0.019; and IgA anti-ALDH4A1: p=0.879). Furthermore, these mediators were also independently associated with CVD in univariate and multivariate models in established RA (Supplementary Table 7). Adjustment by treatment usage in this group did not change these findings.

**Figure 2:**
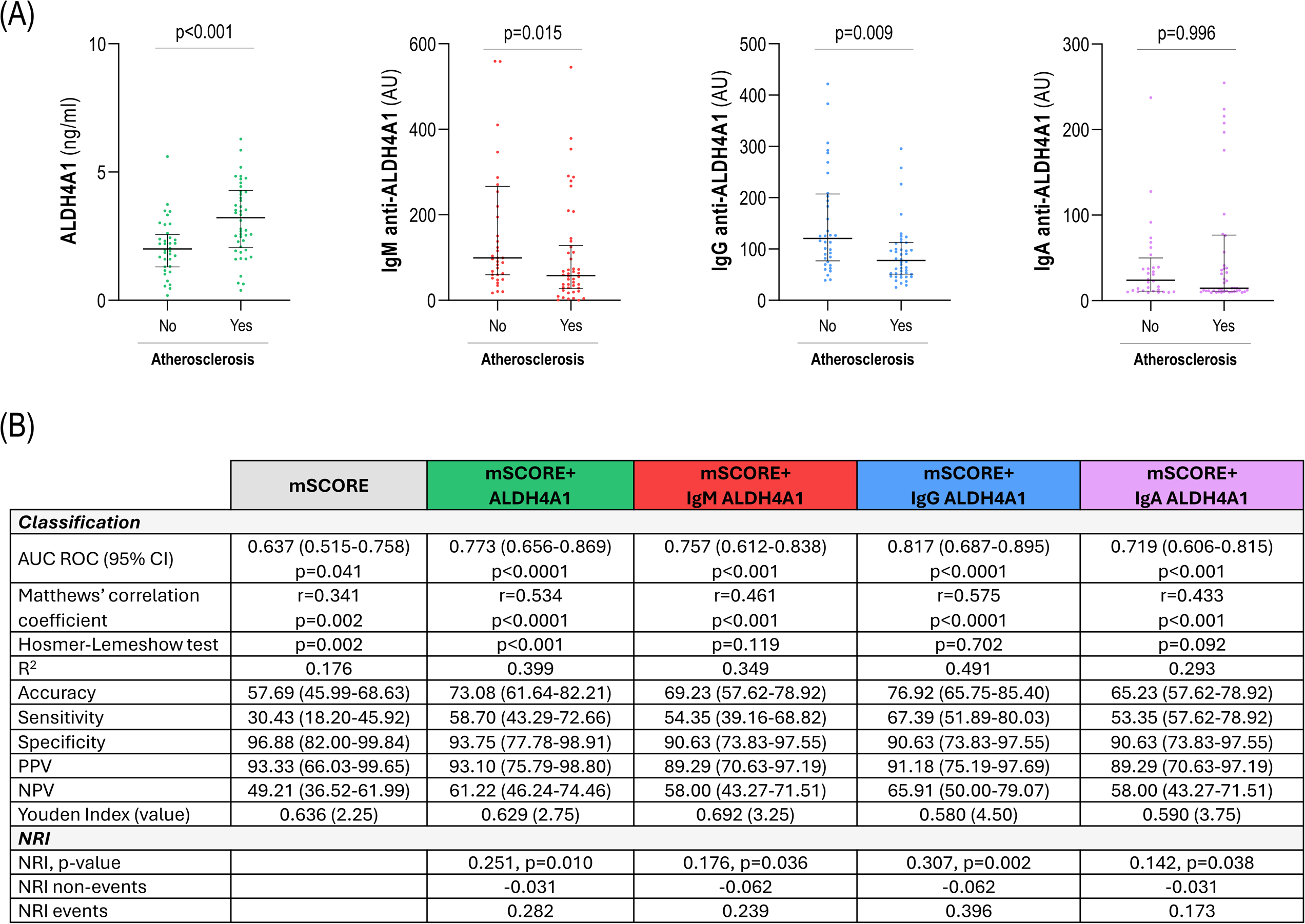
ALDH4A1 and anti-ALDH4A1 serum levels and atherosclerosis in early RA. (A) ALDH4A1 and anti-ALDH4A1 serum levels in early RA patients according to atherosclerosis status. Each dot represents one individual. Bars represent 25th percentile (lower), median and 75th percentile (upper). Differences were assessed by Mann-Withney U tests. P-values are indicated. (B) Classification analyses for the discrimination of atherosclerosis status using the mSCORE alone or in combination with ALDH4A1 or anti-ALDH4A1 antibody levels. Classification, diagnostic performance, calibration metrics, goodness-of-fit statistics and reclassification measures were calculated to assess added value.

ALDH4A1, IgM and IgG anti-ALDH4A1 antibody levels alone were able to discriminate between patients with and without atherosclerosis (AUC [95% CI]: 0.718 [0.604-0.831], p<0.001; 0.664 [0.551-0.764], p=0.015; 0.673 [0.551-0.764], p=0.009; respectively). Moreover, adding ALDH4A1 levels to the mSCORE improved the identification of RA patients with atherosclerosis, based on diagnostic and classification metrics (Figure 2B). A similar picture was observed for IgG anti-ALDH4A1 antibodies, whereas a modest effect was retrieved for the IgM isotype, and a marginal, non-significant effect was noted for IgA (Figure 2B). Interestingly, ALDH4A1 or IgG anti-ALDH4A1 resulted in a better discrimination capacity over mSCORE alone (difference between areas = 0.137 [0.031-0.241], p=0.010; and 0.180 [0.071-0.288], p=0.001, respectively) compared to IgM (0.121 [0.003-0.238], p=0.043) or IgA (0.082 [-0.018-0.183], p=0.109). Moreover, although reaching similar Youden indices, the optimal cut-off value achieved by adding IgG antibodies was more realistic for stratification than that of mSCORE alone, ALDH4A1 or the rest of isotypes (Figure 2B), which were mostly specificity-skewed. Finally, NRI analysis confirmed a better reclassification for IgG antibodies, with a larger effect in patients presenting atherosclerosis and a negligible effect in those without.

All these findings demonstrate that ALDH4A1 and anti-ALDH4A1 antibodies were independently associated with atherosclerosis burden in the earliest phases of arthritis. ALDH4A1 and IgG anti-ALDH4A1 antibodies improved risk stratification over conventional algorithms alone, with a better outcome observed for the latter.

### 3. ALDH4A1/anti-ALDH4A1 axis under TNF blockade

Next, the effect of TNF blockade on ALDH4A1 and anti-ALDH4A1 serum levels was evaluated in a group of biological-naïve RA patients prospectively followed-up for 3 months (Supplementary Table 8).

TNF blockade did not lead to changes in ALDH4A1 nor in antibody levels (Figure 3A). However, IgG antibodies were found to be increased upon treatment were observed in good responders (Figure 3B), whereas no effect was found in those with poor outcome. A similar, although non-significant, effect was noted for IgM (p=0.058), whereas no changes were observed in their IgA counterparts. Upon TNF blockade, the change in ALDH4A1 serum levels paralleled that of TNF, but not with clinical improvement (Figure 3C). Interestingly, change in ALDH4A1 serum levels showed a trend with that of IgG anti-ALDH4A1 antibodies (r=0.538, p=0.058), which reached significance in responders (r=0.904, p=0.035; n=5) while being absent in those with poor therapeutic outcome (r=0.381, p=0.352; n=8). No effects were observed with the rest of isotypes.

**Figure 3:**
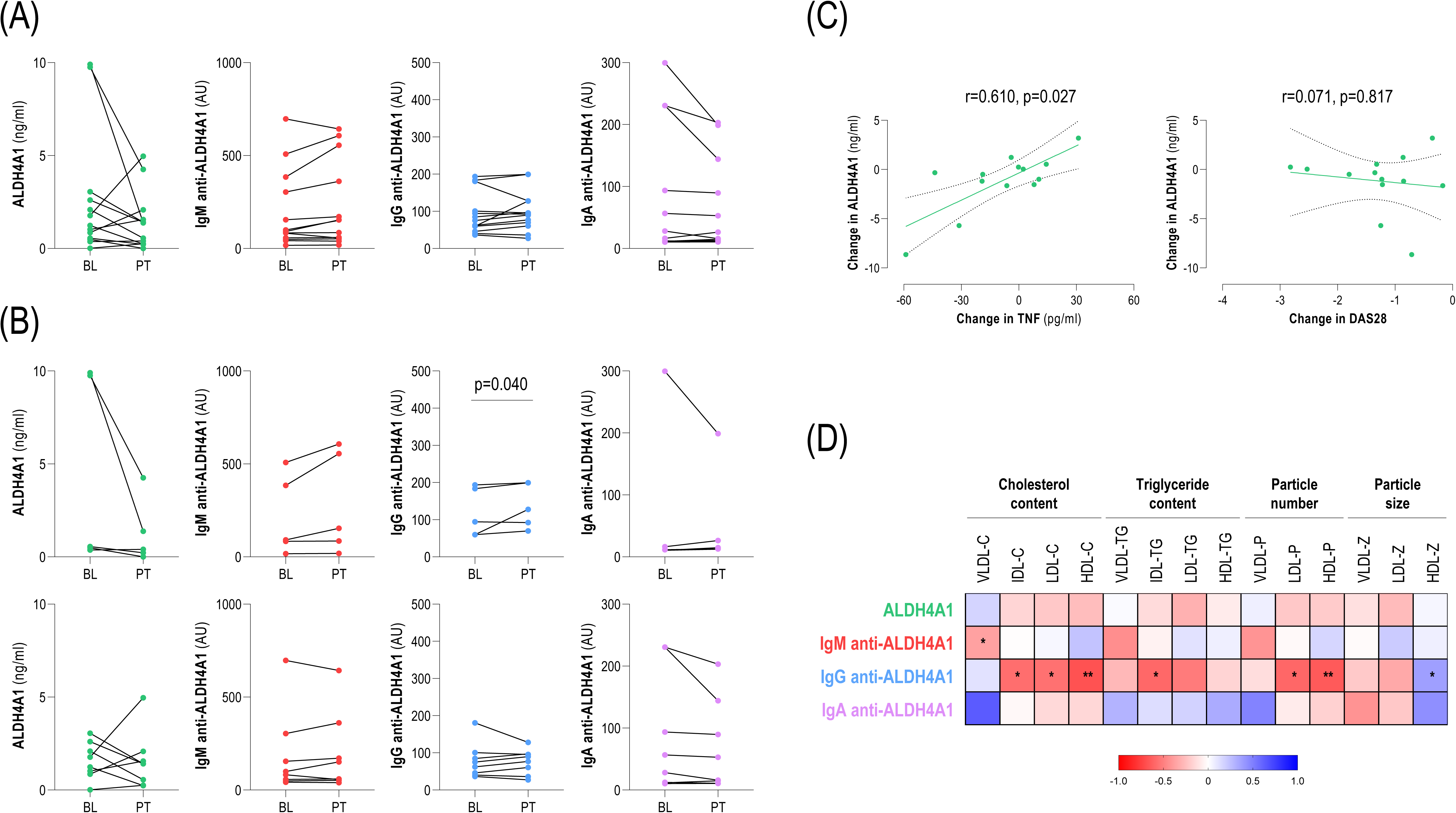
ALDH4A1 and anti-ALDH4A1 serum levels upon TNF blockade in RA. (A) ALDH4A1 and anti-ALDH4A1 antibody serum levels were measured at baseline (BL) and 3-months post-treatment (PT) in biological-naïve RA patients upon in TNF blockade initiation in a prospective study. (B) Patients were stratified according to therapeutic outcomes into those exhibiting good response (n=5, top row) and moderate oor responses (n=8, bottom row). Differences were assessed by paired Wilcoxon tests. Only significant (p<0.050) p-values are shown. (C) Associations between changes in ALDH4A1 serum levels and those of TNF or DAS28 upon TNF blockade. Correlations were assessed by Spearman rank’s test. (D) Matrix depicting the correlations between the changes in lipoprotein profiles (including particle content, number and size) (columns) and those of ALDH4A1 or anti-ALDH4A1 serum levels (rows). Each tile represents a correlation coefficient. Colour gradient varied from blue (positive correlations) to red (negative correlations) according to scale (bottom). The statistically significant p-values from the correlation analyses are indicated as follows: * p<0.050, and ** p<0.010.

Finally, changes in IgG anti-ALDH4A1 serum levels paralleled those of lipoprotein features upon TNF blockade, especially cholesterol content and particle number (Figure 3D). Nevertheless, no associations were retrieved for the rest of isotypes or ALDH4A1 serum levels. Interestingly, these associations mirrored those observed in the cross-sectional analyses (Table 2).

All these results point to subtle changes within the ALDH4A1/anti-ALDH4A1 axis in RA patients upon TNF blockade. These changes connected to the levels of TNF and lipoprotein profiles, and may differ depending on therapeutic outcomes.

## DISCUSSION

The contribution of humoral responses to atherosclerosis in RA remain elusive. Recent research suggests the involvement of mitochondrial-derived antigens in pre-clinical models. Herein we identify ALDH4A1 as a novel antigen relevant to atherosclerosis in early RA. To our knowledge, this is the first description on the role of ALDH4A1 as an antigen in rheumatic and musculoskeletal diseases (RMDs), and the first characterization of anti-ALDH4A1 responses in human patients.

Our findings highlight a relevant role of ALDH4A1 as a novel player and biomarker of atherosclerosis in RA. Despite not elevated across the whole RA group, markedly increased ALDH4A1 levels were observed in patients with atherosclerosis or prior CVD. Aldh4a1 has been described as a protective oxidative stress-induced gene (24). Current literature is supportive of sustained ROS and oxidative stress in RA (25), which may account for this elevation. Enhanced apoptosis, altered mitochondrial clearance and dysfunction (26), or defective efferocytosis (27) may also contribute, consistent with the apoptosis-related pathways and cfDNA associations detected in our analyses. Furthermore, associations with lipoprotein profiles and oxylipin species in early RA were found. Experimental data linked ALDH4A1 to lipid metabolism via Nrf2 (28), aligning with its hepatic expression in atherosclerosis animal models (5). Evidence from our group and others have also demonstrated a link between oxidative stress, impaired lipoprotein levels and CVD (29). Oxidative stress has been also linked to immune responses at the liver via maladaptive responses and neoepitope formation (30), which may relate to our findings on urate/AAT levels and contribute to antibody generation against ALDH4A1, respectively. Furthermore, ALDH4A1 neutralization was associated with profound changes in lipid pathways (5), including eicosanoid and arachidonic metabolism. Taken together, our lipidomic and proteomic approaches proved a robust connection with lipid and immune pathways functionally related to atherosclerosis. At the cellular level, increased ALDH4A1 may led to disturbances within the proline-glutamate pathway (31). This is consistent with reports linking glutamate dysregulation to both arthritis pathogenesis and atherosclerosis outcomes (32,33), thereby reinforcing these functional links. Altogether, these lines of evidence support the concept that mitochondrial antigens play a central role in autoimmunity and its comorbidities, even though antigenic repertoires remain incompletely characterized.

A major contribution of this study was the comprehensive characterization of the humoral anti-ALDH4A1 responses, across RA stages and antibody isotypes. The identification of antigenic targets in atherosclerosis has been the object of profound research. Since most studies followed a targeted approach, antigens described in the literature belong to endothelial or vascular antigens, including lipoproteins, whereas other components have been neglected. Our findings highlighted the involvement of mitochondrial proteins as relevant antigenic targets. Although antibodies against ALDH4A1 have been found to be protective in mouse models, anti-ALDH4A1 responses remained largely unexplored in humans. Lorenzo et al. (5) found no differences in IgM levels depending on atherosclerosis status in human individuals, whereas our findings suggest a potential protective role. Important differences in age and disease background between populations may account for this discrepancy. Of note, immune mechanisms are known to account for a larger proportion of CV risk in autoimmunity (34,35), which may explain a stronger effect in our cohort. Furthermore, antibody generation may differ between general population and autoimmune patients, due to genetic factors, tolerance breakdown, altered antigen clearance, or posttranslational modifications, which could result in differences within humoral responses. Protective activities of IgM antibodies seem to relate to neutralization of oxidation-specific epitopes, which are enriched within IgM repertoires (36,37). However, this effect is less clear for other specificities (36,38), and few comparative studies are available, especially in autoimmune patients. In addition to that of IgM, our findings went further by demonstrating a protective effect of IgG antibodies. The significance of this observation is two-fold. First, as it challenges the existing dogma of ascribing protective properties to IgM, whereas IgG use to be considered as proatherogenic. Second, our findings underscore the need of considering both antigenic specificity as well as isotype to decipher the effect of antibodies in atherosclerosis.

Comparative analyses demonstrated stronger protective associations for IgG than for IgM anti-ALDH4A1 antibodies in RA. Decreased antibody levels in RA may relate to their protective effect, probably in line with an enhanced consumption. Although IgM antibodies are more efficient in neutralizing antigens, IgG can trigger a wider range of effector functions (39). As these functions are dictated by isotype/FcR engagement, deciphering these interactions will be central to understand their effect. Interestingly, IgG anti-ALDH4A1 antibodies predominantly belonged to the IgG3 subclass, consistent with the murine A12 antibody data (5), hence expanding their relevance to the human setting. Moreover, IgG3 antibodies are equipped with potent neutralization/opsonization and FcR-mediated phagocytic properties (39), which may account for a more protective effect. Moreover, they are the strongest inducers or complement activation among IgG (39), hence supporting our findings with complement levels. Furthermore, IgG3 are among the strongest engagers of FcgRIIB, an inhibitory receptor associated with atherosclerosis protection (40,41). Altogether, this functional profile may explain our findings relative to IgG3, and IgG in general, in our study. Identifying the specific FcR engaged by IgG anti-ALDH4A1 antibodies and their cellular context, including immune and stromal compartments, will be instrumental to better understand protective mechanisms. Overall, increased FcR engagement and/or complement consumption may thus account for the decreased levels observed in patients. Furthermore, our study ruled out a potential effect of IgA anti-ALDH4A1 in atherosclerosis. The role of IgA antibodies in atherosclerosis has received little attention, especially compared to that of IgM or IgG (42). Our data demonstrate that these antibodies can be detected in RA, probably suggesting an ongoing immune response at mucosal sites, either to self-antigen or via molecular mimicry. Due to the role of mucosal stress in RA, these antibodies may likely be involved in disease pathogenesis rather than atherosclerosis occurrence itself. Collectively, our findings point to a progressive diversification of the humoral response against ALDH4A1 along the RA spectrum, as indicated by the changing associations among isotypes across stages. The declining correlation between IgG/M levels may be indicative or disturbed class-switch, aberrant T-cell collaboration, extrafollicular responses, or altered immune regulation. Our associations with APRIL may support this interpretation, as APRIL has been linked to class switch to IgG3 (43), especially in extrafollicular and marginal zone B-cells, both subsets being linked to atherosclerosis (38,44), and suggests the role of TLR activation in this phenomenon (45), which may explain a differential role in RA. Alternatively, ‘natural’ B1 cells have been found to be able to migrate to secondary lymphoid organs and undergo in class-switch to IgG or IgA under certain stimuli (46–48). Further studies combining single-cell technology with deep phenotyping are warranted to gain insight into B-cell dynamics underlying anti-ALDH4A1 responses.

Analyses of the IgG anti-ALDH4A1 responses pointed to associations with favourable lipoprotein, suggesting potential therapeutic implications. Experimental data indicates that antibodies may abrogate the liver-immune crosstalk in mouse models (30). This is in line with our findings on the association between IgG anti-ALDH4A1 and urate or AAT levels, which have been linked with CV risk (49–52). Longitudinal analyses under TNF blockade provided additional insight. First, changes in IgG (and, to a lower extent, IgM) may shed new light into the cardio-protective effect of TNF inhibitors, which had been associated with good therapeutic outcomes (53), although underlying mechanisms remain ill-defined. On the other hand, ALDH4A1 changes correlated with TNF dynamics, thus supporting their link with TNF- and apoptosis-related pathways in our proteomic platform. Furthermore, changes in IgG levels were paralleled with those of lipoprotein features, hence mirroring our cross-sectional analysis and pointing to changes in cholesterol metabolism. These findings also align with evidence from the therapeutic effect of anti-ALDH4A1 antibodies in mice models (5). Notably, changes in lipoprotein levels during inflammation are not well understood in RA (6). Current evidence suggest that inflammation itself plays a smaller role than expected (54), thereby suggesting the participation of other mechanisms. Actually, changes in disease activity do not automatically translate into lipoprotein normalization in patients under DMARD treatment (6), and our results ruled out an association with changes in DAS28. Our group has extensively reported the role of autoantibodies in this scenario (12,55,56). The findings herein presented add to this notion by expanding the antigenic repertoire involved in this phenomenon.

Finally, ALDH4A1 and IgG anti-ALDH4A1 hold promise as biomarkers of CV risk in early RA. Their independent association with atherosclerosis and their ability to enhance risk stratification beyond conventional algorithms, with a substantial effect observed for IgG antibodies, support their potential clinical utility. These findings may cover a major clinical unmet need in this setting (57). Notably, their predictive value appears independent of disease activity, the only nontraditional risk factor currently recognized in current guidelines (57), thus highlighting their added value to guide clinical implementation in the future. Moreover, these findings appeared early along RA course and were independent of disease duration, thereby strengthening previous findings and underscoring the need of early strategies.

In summary, ALDH4A1 and anti-ALDH4A1 antibodies could be considered as biomarkers of CV outcomes along the whole RA spectrum, especially during the earliest stages. From a translational standpoint, these findings bridge molecular and clinical dimensions: elucidating functional pathways, improving risk prediction, and informing therapeutic strategies. ALDH4A1 could represent a mechanistic link between immune responses, lipid disturbances, and cardiovascular risk in RA. Prospective studies are warranted to further dissect isotype-specific prognostic implications and clarify the role of IgA antibodies in disease manifestations beyond atherosclerosis. The ALDH4A1/anti-ALDH4A1 axis hold promise as a central player in atherosclerosis development in autoimmunity. This study paves the ground for a deeper analysis of mitochondrial proteomes in RA, and potentially in RMDs, with a special focus on comorbidities.

## Author contributions

All authors were involved in drafting the manuscript or revising it critically for important intellectual content and all the authors gave their approval of the final version of the manuscript to be published.

Study conception and design: JRC, AS

Acquisition of data: DM-P, MAL, AIPA, RC, SAC, NA, MG, AS, JRC

Analysis and interpretation of data: DM-P, AS, JRC

## Data Availability Statement

The data underlying this article are available in the article and in its online supplementary material.

## Acknowledgements

The authors would like to thank the “Liga Reumatológica Asturiana” for their collaboration and support.

---

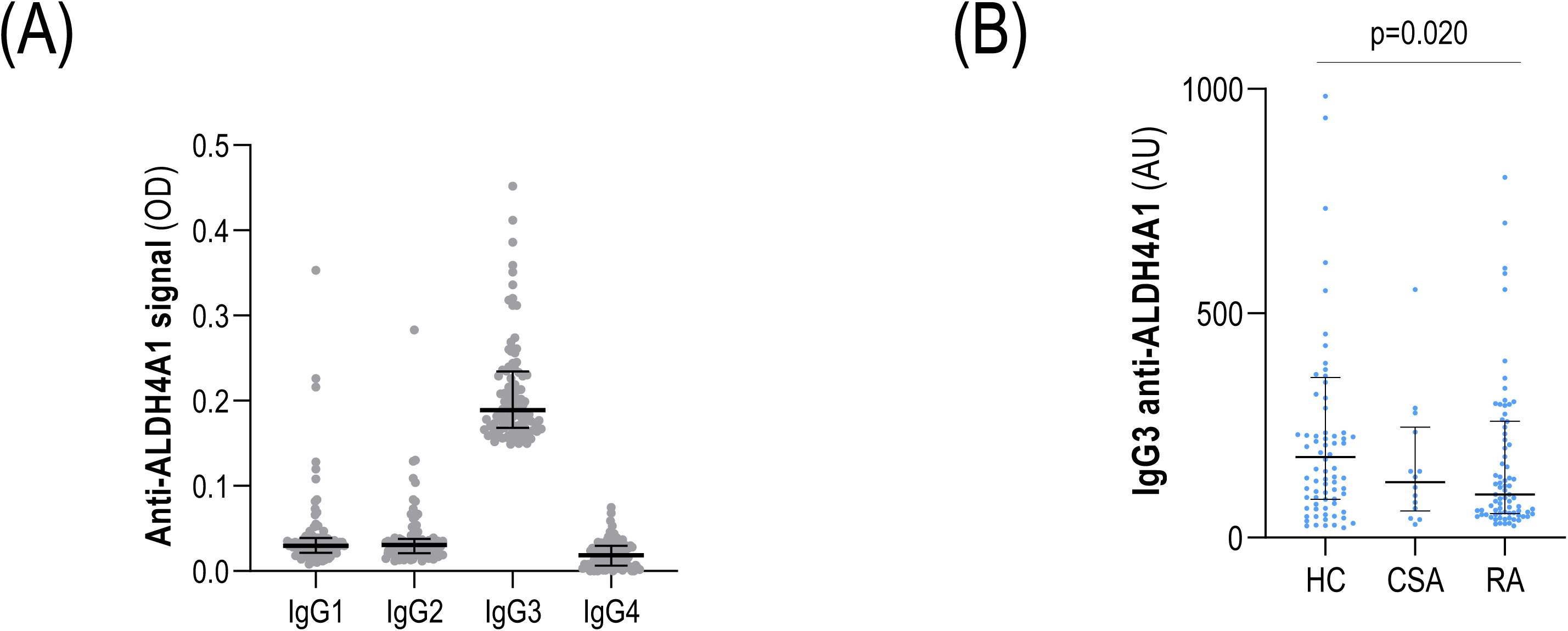

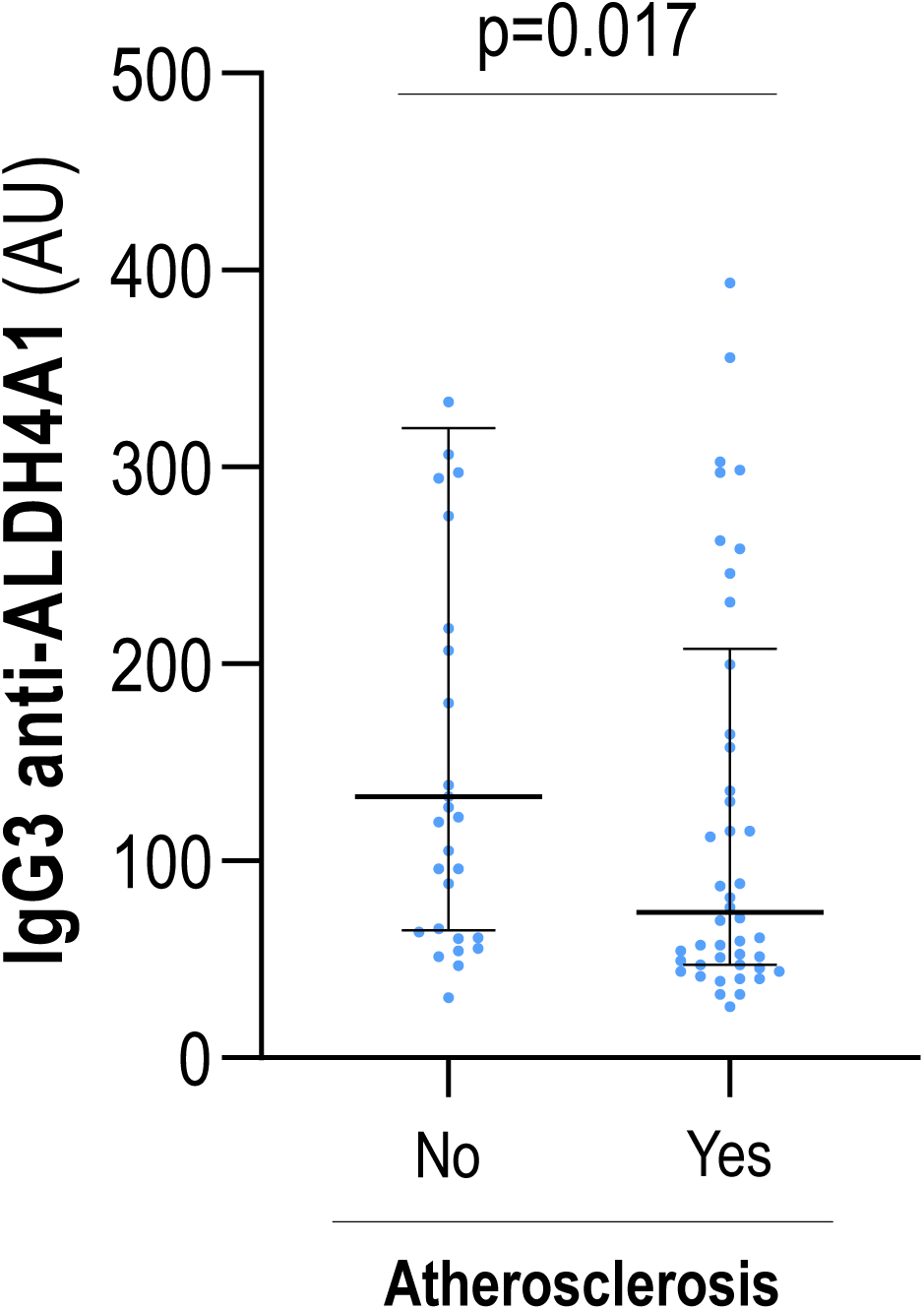

## REFERENCES

1. Avina-Zubieta JA, Thomas J, Sadatsafavi M, Lehman AJ, Lacaille D. Risk of incident cardiovascular events in patients with rheumatoid arthritis: a meta-analysis of observational studies. Ann Rheum Dis 2012;71:1524–1529. Available at: http://ard.bmj.com/lookup/doi/10.1136/annrheumdis-2011-200726.

2. Porsch F, Binder CJ. Autoimmune diseases and atherosclerotic cardiovascular disease. Nat Rev Cardiol 2024;21:780–807.

3. Porsch F, Mallat Z, Binder CJ. Humoral immunity in atherosclerosis and myocardial infarction: from B cells to antibodies. Cardiovasc Res 2021.

4. Raposo-Gutiérrez I, Rodríguez-Ronchel A, Ramiro AR. Atherosclerosis antigens as targets for immunotherapy. Nature Cardiovascular Research 2023;2:1129–1147.

5. Lorenzo C, Delgado P, Busse CE, Sanz-Bravo A, Martos-Folgado I, Bonzon-Kulichenko E, et al. ALDH4A1 is an atherosclerosis auto-antigen targeted by protective antibodies. Nature 2021;589:287–292.

6. Robertson J, Peters MJ, McInnes IB, Sattar N. Changes in lipid levels with inflammation and therapy in RA: a maturing paradigm. Nat Rev Rheumatol 2013;9:513–23. Available at: http://www.ncbi.nlm.nih.gov/pubmed/23774906.

7. Aletaha D, Neogi T, Silman AJ, Funovits J, Felson DT, Bingham CO, et al. 2010 rheumatoid arthritis classification criteria: an American College of Rheumatology/European League Against Rheumatism collaborative initiative. Ann Rheum Dis 2010;69:1580–8. Available at: http://www.ncbi.nlm.nih.gov/pubmed/20699241.

8. Steenbergen HW van, Aletaha D, Beaart-van de Voorde LJJ, Brouwer E, Codreanu C, Combe B, et al. EULAR definition of arthralgia suspicious for progression to rheumatoid arthritis. Ann Rheum Dis 2017;76:491–496. Available at: http://ard.bmj.com/lookup/doi/10.1136/annrheumdis-2016-209846.

9. Rodríguez-Carrio J, Alperi-López M, López P, Ballina-Garciá FJ, Abal F, Suárez A. Antibodies to high-density lipoproteins are associated with inflammation and cardiovascular disease in rheumatoid arthritis patients. Translational Research 2015;166.

10. Smolen JS, Landewé R, Bijlsma J, Burmester G, Chatzidionysiou K, Dougados M, et al. EULAR recommendations for the management of rheumatoid arthritis with synthetic and biological disease-modifying antirheumatic drugs: 2016 update. Ann Rheum Dis 2017;76:960–977. Available at: http://ard.bmj.com/lookup/doi/10.1136/annrheumdis-2016-210715.

11. Gestel AM Van, Anderson JJ, Riel PLCM Van, Boers M, Haagsma CJ, Rich B, et al. ACR and EULAR improvement criteria have comparable validity in rheumatoid arthritis trials. Journal of Rheumatology 1999;26:705–711.

12. Rodríguez-Carrio J, Alperi-López M, López P, Pérez-Álvarez ÁI, Robinson GA, Alonso-Castro S, et al. Humoral responses against HDL are linked to lipoprotein traits, atherosclerosis, inflammation and pathogenic pathways during early arthritis stages. Rheumatology 2023.

13. Touboul P-J, Hennerici MG, Meairs S, Adams H, Amarenco P, Bornstein N, et al. Mannheim carotid intima-media thickness consensus (2004-2006). An update on behalf of the Advisory Board of the 3rd and 4th Watching the Risk Symposium, 13th and 15th European Stroke Conferences, Mannheim, Germany, 2004, and Brussels, Belgium, 2006. Cerebrovasc Dis 2007;23:75–80.

14. O’Rourke MF, Staessen JA, Vlachopoulos C, Duprez D, Plante G e. E. Clinical applications of arterial stiffness; definitions and reference values. Am J Hypertens 2002;15:426–444. Available at: https://academic.oup.com/ajh/article-lookup/doi/10.1016/S0895-7061(01)02319-6.

15. Mallol R, Amigó N, Rodríguez MA, Heras M, Vinaixa M, Plana N, et al. Liposcale: a novel advanced lipoprotein test based on 2D diffusion-ordered 1H NMR spectroscopy. J Lipid Res 2015;56:737–746. Available at: https://linkinghub.elsevier.com/retrieve/pii/S0022227520356017.

16. Rodríguez-Carrio J, Coras R, Alperi-López M, López P, Ulloa C, Ballina-García FJ, et al. Profiling of Serum Oxylipins During the Earliest Stages of Rheumatoid Arthritis. Arthritis & Rheumatology 2021:art.41537. Available at: https://onlinelibrary.wiley.com/doi/10.1002/art.41537.

17. Quehenberger O, Armando AM, Brown AH, Milne SB, Myers DS, Merrill AH, et al. Lipidomics reveals a remarkable diversity of lipids in human plasma. J Lipid Res 2010;51:3299–305. Available at: http://www.ncbi.nlm.nih.gov/pubmed/20671299.

18. Wang Y, Armando AM, Quehenberger O, Yan C, Dennis EA. Comprehensive ultra-performance liquid chromatographic separation and mass spectrometric analysis of eicosanoid metabolites in human samples. J Chromatogr A 2014;1359:60–9. Available at: http://www.ncbi.nlm.nih.gov/pubmed/25074422.

19. Lundberg M, Eriksson A, Tran B, Assarsson E, Fredriksson S. Homogeneous antibody-based proximity extension assays provide sensitive and specific detection of low-abundant proteins in human blood. Nucleic Acids Res 2011;39:e102–e102. Available at: https://academic.oup.com/nar/article-lookup/doi/10.1093/nar/gkr424.

20. Tobío-Parada U, Rodríguez-Carrio J, Martínez-Zapico A, Pérez-Álvarez ÁI, Suárez-Díaz S, Suárez A, et al. Imbalance between serum DNase-I protein levels and enzymatic activity in SLE: link with mitochondrial-DNA and low-density granulocytes. Rheumatology 2025.

21. Szklarczyk D, Gable AL, Nastou KC, Lyon D, Kirsch R, Pyysalo S, et al. The STRING database in 2021: customizable protein–protein networks, and functional characterization of user-uploaded gene/measurement sets. Nucleic Acids Res 2021;49:D605–D612. Available at: https://academic.oup.com/nar/article/49/D1/D605/6006194.

22. Kanehisa M, Furumichi M, Sato Y, Ishiguro-Watanabe M, Tanabe M. KEGG: integrating viruses and cellular organisms. Nucleic Acids Res 2021;49:D545–D551. Available at: https://academic.oup.com/nar/article/49/D1/D545/5943834.

23. Xia J, Wishart DS. Using MetaboAnalyst 3.0 for Comprehensive Metabolomics Data Analysis. Curr Protoc Bioinformatics 2016;55:14.10.1–14.10.91. Available at: http://doi.wiley.com/10.1002/cpbi.11.

24. Yoon K-A, Nakamura Y, Arakawa H. Identification of ALDH4 as a p53-inducible gene and its protective role in cellular stresses. J Hum Genet 2004;49:134–140.

25. Smallwood MJ, Nissim A, Knight AR, Whiteman M, Haigh R, Winyard PG. Oxidative stress in autoimmune rheumatic diseases. Free Radic Biol Med 2018;125:3–14.

26. Weyand CM, Wu B, Huang T, Hu Z, Goronzy JJ. Mitochondria as disease-relevant organelles in rheumatoid arthritis. Clin Exp Immunol 2023;211:208–223.

27. Abdolmaleki F, Farahani N, Gheibi Hayat SM, Pirro M, Bianconi V, Barreto GE, et al. The Role of Efferocytosis in Autoimmune Diseases. Front Immunol 2018;9.

28. Pang S, Lynn DA, Lo JY, Paek J, Curran SP. SKN-1 and Nrf2 couples proline catabolism with lipid metabolism during nutrient deprivation. Nat Commun 2014;5:5048.

29. Rodríguez-Carrio J, Alperi-López M, López P, López-Mejías R, Alonso-Castro S, Abal F, et al. High triglycerides and low high-density lipoprotein cholesterol lipid profile in rheumatoid arthritis: A potential link among inflammation, oxidative status, and dysfunctional high-density lipoprotein. J Clin Lipidol 2017.

30. Busch CJ, Hendrikx T, Weismann D, Jäckel S, Walenbergh SMA, Rendeiro AF, et al. Malondialdehyde epitopes are sterile mediators of hepatic inflammation in hypercholesterolemic mice. Hepatology 2017;65:1181–1195.

31. Srivastava D, Singh RK, Moxley MA, Henzl MT, Becker DF, Tanner JJ. The Three-Dimensional Structural Basis of Type II Hyperprolinemia. J Mol Biol 2012;420:176–189.

32. Lehn-Stefan A, Peter A, Machann J, Schick F, Randrianarisoa E, Heni M, et al. Elevated Circulating Glutamate Is Associated With Subclinical Atherosclerosis Independently of Established Risk Markers: A Cross-Sectional Study. J Clin Endocrinol Metab 2021;106:e982–e989.

33. Flood SL, Duance VC, Mason DJ. The role of glutamate signalling in rheumatoid arthritis. Int J Exp Pathol 2004;85.

34. Gonzalez A, Maradit Kremers H, Crowson CS, Ballman K V, Roger VL, Jacobsen SJ, et al. Do cardiovascular risk factors confer the same risk for cardiovascular outcomes in rheumatoid arthritis patients as in non-rheumatoid arthritis patients? Ann Rheum Dis 2008;67:64–9. Available at: http://www.ncbi.nlm.nih.gov/pubmed/17517756.

35. England BR, Thiele GM, Anderson DR, Mikuls TR. Increased cardiovascular risk in rheumatoid arthritis: mechanisms and implications. BMJ 2018:k1036. Available at: http://www.bmj.com/lookup/doi/10.1136/bmj.k1036.

36. Ransegnola BP, Pattarabanjird T, McNamara CA. Tipping the Scale: Atheroprotective IgM-Producing B Cells in Atherosclerosis. Arterioscler Thromb Vasc Biol 2024;44:1906–1915.

37. Witztum JL, Lichtman AH. The Influence of Innate and Adaptive Immune Responses on Atherosclerosis. Annual Review of Pathology: Mechanisms of Disease 2014;9:73–102.

38. Jones PW, Mallat Z, Nus M. T-Cell/B-Cell Interactions in Atherosclerosis. Arterioscler Thromb Vasc Biol 2024;44:1502–1511.

39. Deroissart J, Binder CJ, Porsch F. Role of Antibodies and Their Specificities in Atherosclerotic Cardiovascular Disease. Arterioscler Thromb Vasc Biol 2024;44:2154–2168.

40. Mendez-Fernandez Y V., Stevenson BG, Diehl CJ, Braun NA, Wade NS, Covarrubias R, et al. The inhibitory FcγRIIb modulates the inflammatory response and influences atherosclerosis in male apoE−/− mice. Atherosclerosis 2011;214:73–80.

41. Bagchi-Chakraborty J, Francis A, Bray T, Masters L, Tsiantoulas D, Nus M, et al. B Cell Fcγ Receptor IIb Modulates Atherosclerosis in Male and Female Mice by Controlling Adaptive Germinal Center and Innate B-1-Cell Responses. Arterioscler Thromb Vasc Biol 2019;39:1379–1389.

42. Taylor JA, Hutchinson MA, Gearhart PJ, Maul RW. Antibodies in action: the role of humoral immunity in the fight against atherosclerosis. Immunity & Ageing 2022;19:59.

43. Stein J V, López-Fraga M, Elustondo FA, Carvalho-Pinto CE, Rodríguez D, Gómez-Caro R, et al. APRIL modulates B and T cell immunity. J Clin Invest 2002;109:1587–98.

44. Harrison J, Newland SA, Jiang W, Giakomidi D, Zhao X, Clement M, et al. Marginal zone B cells produce ‘natural’ atheroprotective IgM antibodies in a T cell–dependent manner. Cardiovasc Res 2024;120:318–328.

45. Hardenberg G, Planelles L, Schwarte CM, Bostelen L van, Huong T Le, Hahne M, et al. Specific TLR ligands regulate APRIL secretion by dendritic cells in a PKR-dependent manner. Eur J Immunol 2007;37:2900–11.

46. Smith FL, Baumgarth N. B-1 cell responses to infections. Curr Opin Immunol 2019;57:23–31.

47. Halperin ST, ’t Hart BA, Luchicchi A, Schenk GJ. The Forgotten Brother: The Innate-like B1 Cell in Multiple Sclerosis. Biomedicines 2022;10:606.

48. Mora JR, Andrian UH von. Differentiation and homing of IgA-secreting cells. Mucosal Immunol 2008;1:96–109.

49. Gagliardi ACM, Miname MH, Santos RD. Uric acid: A marker of increased cardiovascular risk. Atherosclerosis 2009;202:11–17.

50. Yun KE, Shin CY, Yoon YS, Park HS. Elevated alanine aminotransferase levels predict mortality from cardiovascular disease and diabetes in Koreans. Atherosclerosis 2009;205:533–537.

51. Hu X, Liu J, Li W, Wang C, Li G, Zhou Y, et al. Elevated serum uric acid was associated with pre-inflammatory state and impacted the role of HDL-C on carotid atherosclerosis. *Nutrition*, Metabolism and Cardiovascular Diseases 2022;32:1661–1669.

52. Kim S-K, Kim D-J, Kim S-H, Lee Y-K, Park S-W, Cho Y-W, et al. Normal range of alanine aminotransferase concentration is associated with carotid atherosclerosis. Diabetes Res Clin Pract 2010;88:111–116.

53. Dixon WG, Watson KD, Lunt M, Hyrich KL, Silman AJ, Symmons DPM. Reduction in the incidence of myocardial infarction in patients with rheumatoid arthritis who respond to anti–tumor necrosis factor α therapy: Results from the British Society for Rheumatology Biologics Register. Arthritis Rheum 2007;56:2905–2912. Available at: https://onlinelibrary.wiley.com/doi/10.1002/art.22809.

54. Halm VP van, Nielen MMJ, Nurmohamed MT, Schaardenburg D van, Reesink HW, Voskuyl AE, et al. Lipids and inflammation: serial measurements of the lipid profile of blood donors who later developed rheumatoid arthritis. Ann Rheum Dis 2007;66:184–8. Available at: http://www.ncbi.nlm.nih.gov/pubmed/16760255.

55. Rodríguez-Carrio J, Alperi-López M, López-Mejías R, López P, Ballina-García FJ, Abal F, et al. Antibodies to paraoxonase 1 are associated with oxidant status and endothelial activation in rheumatoid arthritis. Clin Sci (Lond) 2016;130:1889–99. Available at: http://www.ncbi.nlm.nih.gov/pubmed/27520507.

56. Rodríguez-Carrio J, López-Mejías R, Alperi-López M, López P, Ballina-García FJ, González-Gay MÁ, et al. Paraoxonase 1 Activity Is Modulated by the rs662 Polymorphism and IgG Anti-High-Density Lipoprotein Antibodies in Patients With Rheumatoid Arthritis: Potential Implications for Cardiovascular Disease. Arthritis & Rheumatology 2016;68:1367–1376.

57. Agca R, Heslinga SC, Rollefstad S, Heslinga M, McInnes IB, Peters MJL, et al. EULAR recommendations for cardiovascular disease risk management in patients with rheumatoid arthritis and other forms of inflammatory joint disorders: 2015/2016 update. Ann Rheum Dis 2017;76:17–28. Available at: https://ard.bmj.com/lookup/doi/10.1136/annrheumdis-2016-209775.

