## Supplementary Tables and figures for "The ALDH4A1/anti-ALDH4A1 axis as a novel player of atherosclerosis in rheumatoid arthritis"

**SUPPLEMENTARY MATERIAL**

**Supplementary Material and methods**

**Supplementary Tables**

**Supplementary Figure legends**

**MATERIAL AND METHODS**

**SUPPLEMENTARY TABLES**

**Supplementary Table 1: Demographic and parameters of established, long-lasting RA (LRA).** Demographic and clinical features of established RA patients recruited as a validation cohort. Variables were expressed as median (interquartile range), mean±SD or n(%), unless otherwise stated, according to the distribution of the variables.

|  | **LRA  n=90** |
| --- | --- |
| Age (years), mean (range) | 53.63  (22.00-87.00) |
| Gender (female/male) | 74/16 |
| ***Clinical features*** |  |
| Disease duration, median (range) (years) | 5.66 (0.50 – 22.83) |
| Age at diagnosis (years), mean (range) | 46.85 (18.00 – 80.92) |
| Tender joint count | 2.00 (5.50) |
| Swollen joint count | 1.00 (3.00) |
| Patient global assessment (VAS 0-100) | 40.00 (40.75) |
| Pain assessment (VAS 0-10) | 4.00 (3.38) |
| DAS28 | 3.58 (1.82) |
| HAQ | 0.87 (1.14) |
| RF+, n(%) | 49 (54.4) |
| ACPA+, n(%) | 57 (63.3) |
| History of CV events, n(%) | 16 (17.7) |
| ***Laboratory parameters*** |  |
| ESR (mm/h) | 17.00 (23.00) |
| CRP (mg/dl) | 1.00 (4.00) |
| AAT (U/l) | 95.00 (14.50) |
| ***Traditional CV risk factors*** |  |
| Hypertension, n(%) | 33 (36.6) |
| Diabetes, n(%) | 7 (7.7) |
| Dyslipidemia, n(%) | 27 (30.0) |
| Smoking, n(%) | 27 (30.0) |
| Obesity (BMI>30 kg/m^2^), n(%) | 16 (17.7) |
| ***Treatments, n(%)*** |  |
| Glucocorticoids | 54 (60.0) |
| Methotrexate | 72 (80.0) |
| TNF blockers | 42 (46.6) |
| IL-6 blockers | 9 (10.0) |
| Leflunomide | 11 (12.0) |

**Supplementary Table 2: ALDH4A1 and anti-ALDH4A1 serum levels and demographic and clinical features in early RA.** Associations between ALDH4A1 or anti-ALDH4A1 serum levels and demographic parameters, clinical features, or traditional CV risk factors and were assessed were analysed by Spearman’s rank tests or Mann-Withney U tests in RA patients. Coefficients (r) and p-values, or p-values for the difference between groups are shown. Those reaching statistical significance were highlighted in bold.

|  | **ALDH4A1** | **IgM anti-ALDH4A1** | **IgG anti-ALDH4A1** | **IgA anti-ALDH4A1** |
| --- | --- | --- | --- | --- |
| Age | **r=0.306**  **p=0.005** | r=-0.169  p=0.130 | r=-0.114  p=0.310 | r=0.009  p=0.937 |
| Gender | p=0.981 | p=0.726 | p=0.699 | p=0.393 |
| ***Clinical features*** |  |  |  |  |
| Duration of symptoms | r=-0.080  p=0.490 | r=-0.009  p=0.940 | r=0.151  p=0.194 | r=0.055  p=0.635 |
| Morning stiffness | r=0.151  p=0.175 | r=0.148  p=0.185 | r=0.131  p=0.239 | r=0.147  p=0.187 |
| Tender joint count | r=0.024  p=0.828 | r=0.076  p=0.496 | r=0.140  p=0.210 | r=0.103  p=0.357 |
| Swollen joint count | r=0.065  p=0.561 | r=0.023  p=0.838 | r=0.097  p=0.384 | r=0.072  p=0.518 |
| Patient global assessment | r=0.078  p=0.491 | r=-0.009  p=0.940 | r=-0.011  p=0.920 | r=0.210  p=0.060 |
| Pain assessment | r=0.044  p=0.696 | r=-0.082  p=0.468 | r=-0.117  p=0.300 | r=0.074  p=0.511 |
| DAS28 | r=0.082  p=0.468 | r=0.078  p=0.489 | r=0.104  p=0.357 | r=0.092  p=0.413 |
| HAQ | r=0.209  p=0.062 | r=0.014  p=0.899 | r=-0.112  p=0.324 | r=0.128  p=0.259 |
| RF | p=0.976 | p=0.519 | p=0.622 | p=0.782 |
| ACPA | p=0.796 | p=0.956 | p=0.675 | p=0.820 |
| ***Laboratory parameters*** |  |  |  |  |
| ESR | r=0.139  p=0.211 | r=0.073  p=0.515 | r=0.099  p=0.374 | r=-0.016  p=0.890 |
| CRP | r=0.145  p=0.192 | r=0.103  p=0.355 | r=0.011  p=0.921 | r=0.079  p=0.479 |
| C3 | r=-0.011  p=0.925 | r=-0.072  p=0.538 | **r=-0.297**  **p=0.010** | r=-0.101  p=0.390 |
| C4 | r=-0.127  p=0.277 | r=-0.053  p=0.652 | **r=-0.237**  **p=0.040** | r=-0.122  p=0.297 |
| Glucose | r=-0.011  p=0.920 | r=-0.214  p=0.065 | r=-0.010  p=0.928 | r=0.155  p=0.165 |
| Urate | r=-0.088  p=0.445 | **r=-0.247**  **p=0.030** | **r=-0.256**  **p=0.016** | r=-0.037  p=0.752 |
| Alanine aminotransferase | r=-0.129  p=0.206 | r=-0.032  p=0.778 | **r=-0.288**  **p=0.010** | r=-0.130  p=0.256 |
| ***Traditional CV risk factors*** |  |  |  |  |
| Hypertension | p=0.253 | p=0.064 | p=0.121 | p=0.591 |
| Diabetes | p=0.091 | **p=0.018** | p=0.830 | p=0.761 |
| Dyslipidaemia | p=0.896 | p=0.058 | **p=0.024** | p=0.057 |
| Smoking | p=0.730 | p=0.366 | p=0.573 | p=0.088 |
| Obesity | p=0.712 | p=0.996 | **p=0.003** | p=0.610 |

**Supplementary Table 3: Associations between ALDH4A1 and anti-ALDH4A1 antibody levels and serum cytokines.** Associations between ALDH4A1 or anti-ALDH4A1 serum levels and those of serum cytokines were analysed by Spearman’s rank tests in RA and CSA groups. Coefficients (r) and p-values are shown. Those reaching statistical significance were highlighted in bold.

|  | **RA** | | | | **CSA** | | | |
| --- | --- | --- | --- | --- | --- | --- | --- | --- |
|  | **ALDH4A1** | **IgM anti-ALDH4A1** | **IgG anti-ALDH4A1** | **IgA anti-ALDH4A1** | **ALDH4A1** | **IgM anti-ALDH4A1** | **IgG anti-ALDH4A1** | **IgA anti-ALDH4A1** |
| IFNa | r=-0.117  p=0.295 | r=0.068  p=0.544 | r=0.067  p=0.551 | r=-0.039  p=0.729 | r=0.348  p=0.222 | r=-0.410  p=0.146 | r=-0.388  p=0.171 | r=0.227  p=0.435 |
| IL-6 | r=-0.043  p=0.699 | r=0.140  p=0.209 | r=0.215  p=0.053 | r=-0.086  p=0.441 | r=0.276  p=0.340 | r=-0.449  p=0.107 | r=-0.476  p=0.086 | r=0.171  p=0.559 |
| TNF | r=-0.052  p=0.641 | r=0.016  p=0.886 | r=-0.044  p=0.698 | r=-0.138  p=0.215 | r=0.077  p=0.793 | r=0.029  p=0.923 | r=-0.073  p=0.805 | r=0.351  p=0.218 |
| IFNg | r=-0.113  p=0.313 | r=0.066  p=0.554 | r=0.052  p=0.643 | r=-0.067  p=0.548 | r=-0.018  p=0.951 | r=-0.061  p=0.837 | r=-0.452  p=0.105 | r=0.279  p=0.334 |
| IL-1b | r=-0.160  p=0.151 | r=-0.010  p=0.928 | r=0.003  p=0.980 | r=-0.085  p=0.449 | **r=0.671**  **p=0.009** | r=-0.278  p=0.336 | r=0.178  p=0.543 | r=0.319  p=0.266 |
| IL-23 | r=-0.110  p=0.327 | r=0.141  p=0.207 | r=0.050  p=0.655 | r=-0.160  p=0.150 | r=-0.128  p=0.663 | r=0.282  p=0.329 | r=0.372  p=0.190 | r=0.430  p=0.125 |
| IL-12 | r=-0.096  p=0.389 | r=0.180  p=0.106 | r=0.087  p=0.439 | r=-0.080  p=0.477 | r=0.158  p=0.590 | r=0.009  p=0.976 | r=0.064  p=0.828 | r=0.195  p=0.503 |
| IL-33 | r=-0.081  p=0.471 | r=0.054  p=0.631 | r=0.036  p=0.749 | r=-0.076  p=0.497 | r=0.185  p=0.527 | r=-0.405  p=0.151 | r=-0.361  p=0.205 | r=0.389  p=0.170 |
| IL-10 | r=-0.134  p=0.231 | r=0.123  p=0.271 | r=0.142  p=0.202 | r=-0.021  p=0.853 | r=0.178  p=0.542 | r=-0.189  p=0.517 | r=0.040  p=0.893 | r=0.117  p=0.691 |
| IL-17 | r=-0.071  p=0.528 | r=0.170  p=0.126 | r=0.184  p=0.097 | r=0.075  p=0.505 | r=-0.251  p=0.387 | **r=0.538**  **p=0.047** | r=0.424  p=0.131 | r=0.517  p=0.058 |
| IL-8 | r=-0.012  p=0.917 | r=0.192  p=0.084 | **r=0.242**  **p=0.029** | r=0.176  p=0.114 | r=0.412  p=0.143 | r=-0.482  p=0.081 | r=-0.244  p=0.400 | r=-0.062  p=0.834 |
| IL-18 | r=0.196  p=0.078 | r=-0.063  p=0.572 | r=-0.022  p=0.847 | r=0.032  p=0.774 | r=0.403  p=0.153 | r=-0.350  p=0.220 | r=-0.389  p=0.169 | r=-0.123  p=0.675 |
| APRIL | r=0.077  p=0.565 | **r=-0.302**  **p=0.021** | r=0.172  p=0.592 | r=-0.155  p=0.245 | r=0.455  p=0.160 | r=-0.518  p=0.102 | r=-0.327  p=0.326 | r=0.073  p=0.831 |
| BAFF | r=-0.028  p=0.837 | r=0.027  p=0.839 | r=-0.114  p=0.395 | r=0.010  p=0.938 | r=0.521  p=0.101 | r=-0.470  p=0.144 | r=-0.014  p=0.968 | r=-0.082  p=0.810 |

**Supplementary Table 4: Pathways derived from oxylipins associated with ALDH4A1.** Oxylipin species found to be associated with ALDH4A1 serum levels (n=12) were entered into a pathway enrichment analysis in MetaboAnalyst using RaMP-DB database. Results were summarized in the table.

|  | **Total** | **Hits** | **p-value (Holm)** | **FDR** | **Enrichment ratio** |
| --- | --- | --- | --- | --- | --- |
| Arachidonic acid (AA, ARA) oxylipin metabolism | 78 | 6 | 1.3·10^-8^ | 4.29·10^-5^ | 30 |
| Metabolism of alpha-linolenic acid | 20 | 4 | 1.12·10^-7^ | 3.71·10^-4^ | 77.8 |
| Eicosanoid metabolism via lipooxygenases (LOX) | 32 | 4 | 8.18·10^-7^ | 0.00271 | 48.6 |
| Docosahexaenoic acid oxylipin metabolism | 35 | 4 | 1.19·10^-6^ | 0.00393 | 44.9 |

**Supplementary Table 5: Subclinical CVD burden in early arthritis groups.** The prevalence of subclinical CVD, namely atherosclerosis burden and vascular stiffness, was assessed by doppler ultrasound following international recommendations in RA and CSA groups. Variables were expressed as median (interquartile range), mean±SD or n(%), unless otherwise stated.

|  | **RA** | **CSA** |
| --- | --- | --- |
| ***Subclinical atherosclerosis*** | n=77 | n=13 |
| Plaque presence, n(%) | 46 (59.7) | 4 (30.7) |
| Plaque number, mean (range) | 0.96 (0-4) | 0.46 (0-3) |
| High-risk plaque, n(%) | 20 (25.9) | 0 (0.0) |
| cIMT (mm) | 0.67±0.10 | 0.58±0.15 |
| ***Vascular stiffness*** | n=77 | n=13 |
| VS | 0.10±0.04 | 0.10±0.03 |
| VD | 0.0044±0.002 | 0.0055±0.0031 |
| VSf | 540.04±239.61 | 504.24±286.37 |
| PSEM | 3.83±1.68 | 3.52±2.00 |

**Supplementary Table 6: Associations between ALDH4A1 and anti-ALDH4A1 antibodies and subclinical CVD features.** Associations between levels of ALDH4A1 or anti-ALDH4A1 and subclinical CVD features were analyzed by Spearman ranks’ tests or Mann-Withney U tests in RA and CSA groups. Coefficients (r) and p-values, or p-values for the difference between groups are shown. Those reaching statistical significance were highlighted in bold.

|  | **RA** | | | | **CSA** | | | |
| --- | --- | --- | --- | --- | --- | --- | --- | --- |
|  | **ALDH4A1** | **IgM anti-ALDH4A1** | **IgG anti-ALDH4A1** | **IgA anti-ALDH4A1** | **ALDH4A1** | **IgM anti-ALDH4A1** | **IgG anti-ALDH4A1** | **IgA anti-ALDH4A1** |
| ***Subclinical atherosclerosis*** | | |  |  |  |  |  |  |
| Plaque presence | **p<0.001** | **p=0.015** | **p=0.009** | p=0.996 | **p=0.020** | p=0.199 | p=0.940 | p=0.106 |
| Plaque number | **r=0.392**  **p<0.001** | **r=-0.214**  **p=0.059** | **r=-0.282**  **p=0.012** | r=0.075  p=0.516 | **r=0.691**  **p=0.009** | r=-0.406  p=0.169 | r=-0.014  p=0.965 | r=0.487  p=0.091 |
| Plaque risk | p=0.919 | p=0.496 | **p=0.061** | p=0.753 | - | - | - | - |
| cIMT | r=0.143  p=0.214 | **r=-0.252**  **p=0.027** | r=-0.212  p=0.064 | r=-0.099  p=0.391 | r=0.182  p=0.552 | r=-0.437  p=0.135 | r=-0.385  p=0.194 | r=-0.063  p=0.837 |
| ***Vascular stiffness*** | |  |  |  |  |  |  |  |
| VS | r=-0.077  p=0.640 | r=0.129  p=0.434 | r=-0.021  p=0.900 | r=-0.171  p=0.298 | r=0.269  p=0.423 | r=-0.237  p=0.483 | r=-0.150  p=0.659 | r=0.155  p=0.649 |
| VD | r=-0.278  p=0.112 | r=0.125  p=0.480 | r=0.084  p=0.637 | r=-0.158  p=0.371 | r=-0.234  p=0.544 | r=0.283  p=0.460 | r=0.317  p=0.406 | r=0.036  p=0.920 |
| VSf | r=0.259  p=0.140 | r=-0.183  p=0.301 | r=-0.144  p=0.415 | r=0.134  p=0.451 | r=0.234  p=0.544 | r=-0.283  p=0.460 | r=-0.317  p=0.406 | r=0.314  p=0.346 |
| PSEM | r=0.278  p=0.112 | r=-0.125  p=0.480 | r=-0.084  p=0.637 | r=0.158  p=0.371 | r=0.234  p=0.544 | r=-0.283  p=0.460 | r=-0.317  p=0.406 | r=-0.042  p=0.907 |

**Supplementary Table 7: ALDH4A1 and anti-ALDH4A1 antibodies as predictors of atherosclerosis.** The role of ALDH4A1 or anti-ALDH4A1 antibody levels as predictors of atherosclerosis occurrence was analyzed by univariate and multivariate logistic regression models in LRA patients. Model 1 represents a univariate model. Model 2 represents a multivariate model adjusted for traditional CV risk factors (age, sex, hypertension, dyslipidaemia, diabetes, smoking, and obesity). Model 3 represents a multivariate model adjusted for traditional CV risk factors and clinical features (model 2 + disease activity, RF, ACPA positivity, and disease duration). Models were run independently for each predictor. Odds ratios (OR), 95% confidence intervals and p-values were calculated for each predictor and model. Those reaching statistical significance were highlighted in bold.

|  | **Model 1** | | **Model 2** | | **Model 3** | |
| --- | --- | --- | --- | --- | --- | --- |
|  | ***OR [95% CI]*** | ***p-value*** | ***OR [95% CI]*** | ***p-value*** | ***OR [95% CI]*** | ***p-value*** |
| ALDH4A1 | 1.253 [1.052 – 1.493] | **0.011** | 1.282 [0.682 – 3.881] | **0.021** | 1.351 [1.047 – 1.743] | **0.021** |
| IgM anti-ALDH4A1 | 0.216 [0.082 – 0.573] | **0.004** | 0.155 [0.041 – 0.584] | **0.006** | 0.739 [0.540 – 1.024] | 0.067 |
| IgG anti-ALDH4A1 | 0.280 [0.119 – 0.659] | **0.008** | 0.220 [0.066 – 0.731] | **0.013** | 0.157 [0.035 – 0.704] | **0.016** |
| IgA anti-ALDH4A1 | 0.991 [0.872 – 1.127] | 0.892 | 0.916 [0.766 – 1.094] | 0.331 | 1.154 [0.821 – 1.622] | 0.409 |

**Supplementary Table 8: Demographic and clinical features of patients undergoing TNF blockade.** Demographic, clinical features and therapeutic outcomes (at 3 months) of biological-naïve RA patients undergoing TNF blockade were summarized. Variables were expressed as median (interquartile range), mean±SD or n(%), unless otherwise stated.

|  | **Biological-naïve RA n=13** | |
| --- | --- | --- |
| Age (years), mean (range) | 46.71  (30.75-65.42) | |
| Gender (female/male) | 12/1 | |
| ***Clinical features*** |  |  |
| Disease duration, median (range) (years) | 1.54 (1.00 – 7.17) | |
| Age at diagnosis (years), mean (range) | 44.28 (29.08 – 62.75) | |
| RF+, n(%) | 5 (38.4) | |
| ACPA+, n(%) | 6 (46.1) | |
| ***Therapeutic outcomes*** | *Baseline (BL)* | *Post-treatment (PT)* |
| Tender joint count | 9.00 (5.00) | 4.50 (3.00) |
| Swollen joint count | 5.00 (3.00) | 3.00 (1.69) |
| ESR (mm/h) | 13.00 (28.00) | 9.5 (16.25) |
| CRP (mg/dl) | 3.00 (4.15) | 1.00 (1.50) |
| Patient global assessment (VAS 0-100) | 66.00 (17.00) | 35.00 (20.28) |
| Pain assessment (VAS 0-10) | 6.00 (2.90) | 3.25 (2.97) |
| DAS28 | 5.15 (1.99) | 3.80 (1.97) |
| HAQ | 1.20 (0.79) | 0.87 (0.85) |

**SUPPLEMENTARY FIGURE LEGENDS**

**Supplementary Figure 1: Analysis of IgG anti-ALDH4A1 responses.** (A) Subclass analysis revealed that IgG3 subclass predominated within IgG anti-ALDH4A1 responses in our study. OD for each sample, after subtracting background, was shown. (B) IgG3 anti-ALDH4A1 serum levels (in AU) were calculated from standard curves (pooled sera). Differences across groups mirrored those obtained with total IgG responses (Figure 1). Each dot represents one individual. Bars represent 25th percentile (lower), median and 75th percentile (upper). Differences were assessed by Kruskal-Wallis tests with Dunn-Bonferroni post-hoc tests. The p-values from the latter were indicated.

**Supplementary Figure 2: IgG3 anti-ALDH4A1 and atherosclerosis in RA.** IgG3 anti-ALDH4A1 serum levels (AU) in RA patients according to the atherosclerosis status were shown. Differences between groups mirrored those obtained with total IgG responses (Figure 2). Each dot represents one individual. Bars represent 25th percentile (lower), median and 75th percentile (upper). Differences were assessed by Mann-Withney U tests. The p-value from the latter was indicated.
